# Estimating the Effects of Individual Nurses on ICU Outcomes

**DOI:** 10.1101/2020.11.04.20226340

**Authors:** Stephen E. Brossette, Ning Zheng, Daisy Y. Wong, Patrick A. Hymel

## Abstract

A better understanding of the effects of nursing on clinical outcomes could be used to improve the safety, efficacy, and efficiency of inpatient care. However, measuring the performance of individual nurses is complicated by the non-random assignment of nurses to patients, a process that is confounded by unobserved patient, management, workforce, and institutional factors. Using the MIMIC-III ICU database, we estimate the effects of individual registered nurses (RNs) on the probability of acute kidney injury (AKI) in the ICU. We control for significant unobserved heterogeneity by exploiting panel data with 12-hour fixed effects, and use a linear probability model to estimate the near-term marginal effects of individual RN assignments. Among 270 ICU RNs, we find 15 excess high-side outliers, and 4 excess low-side outliers. We estimate that in one twelve-hour work shift, each high-side RN outlier increases the probability of AKI by about 4 percentage points, and in 25 work shifts, causes about one additional AKI. Conversely, each low-side outlier prevents about one AKI in 50 work shifts. Given the fine-grained nature of the fixed effects employed, we believe that the estimated individual nursing effects are approximately causal. We discuss our contribution to the literature and identify potential use cases for clinical deployment.

## Introduction

Efforts to quantify the value of nursing often focus on the effects of staffing intensity, nurse education, and nurse experience (McHugh et al, 2013; Lake et al, 2012; Dall et al, 2009; Blegen, 2013; Dabney 2015; Konetzka 2007; Kutney-Lee 2015; Mchugh 2013; Needleman 2002; Weiss 2011; Yakusheva 2014). Without designed or natural experiments, however, unbiased estimates of nursing effects have been difficult to obtain, and work to estimate the effects of individual nurses is so far limited to one study (Yakusheva 2014). The fundamental difficulty in estimating nursing performance is that the assignment of a nurse to a patient is not random, and estimates of nursing effects, collectively or individually, are confounded by unobserved factors that are correlated with nursing assignment – things like aggregate workload, nursing availability, patient-level case complexity, prior assignments, and management preferences. In an experimental evaluation of nursing effects, assignment would be randomized like treatment is randomized in a well-designed drug trial. Randomization would ensure that patients assigned to one nurse do not systematically differ from those assigned to others, within chance, and that estimated nursing effects are causal. Without experimental data, however, the challenge is to model observational data and infer causal effects. This is possible if all factors related to nursing assignment are accounted for so that assignment itself is exogenous. However, since many of these factors are difficult to observe, models of observational cross-sectional data likely omit at least some of them, thereby producing biased estimates of nursing effects.

Short of designed experiments where assignment is randomized, methods to account for unobserved, assignment-related factors include exploiting natural experiments, finding instrumental variables, and using unobserved-effects models on panel data. However, except for the use of facility-level fixed effects in models that evaluate aggregate nursing features and staffing levels (Kutney-Lee et al, 2015; Konetzka et al, 2007; Park & Sterns, 2009), we are unable to find examples that use any of these methods. In this work, we use a fixed-effects model on panel data computed from the MIMIC-III ICU database to estimate the effects of individual nurses on the likelihood of acute kidney injury (AKI) in the ICU. By assigning fixed effects to 12-hour substays, we control for significant sources of unobserved heterogeneity. These include individual patient-level factors like illness burden, medical and surgical histories, physician assignments, and care plans. They also include system and location-level features like caseload and nurse staffing. For example, if an RN is assigned patients who are intrinsically more likely to develop AKI, or to patients with physicians that increase AKI risk, the fixed effects incrementally adjust and control for these risks at each substay. If an ICU is busy, and aggregate care demands affect RN assignment, the changing caseload demand is captured by the substay-level fixed effect. The RN assignment algorithm itself, as well as time-sensitive nursing supply factors are also controlled for since they are effectively constant at the substay level. With substay-level fixed effects, the fixed-effects estimator adjusts for factors like these, observed or not, every 12 hours.

We model the effects of individual nurses on acute kidney injury and chose AKI because it is both clinically significant and computable from routine laboratory data (Khwaja, 2012; Mehta et al., 2007). It is also associated with adverse in-hospital events like myocardial infarction and sepsis (Hoste, 2009), and occurs with inappropriate medication, fluid and electrolyte management, in addition to a variety of other pre-renal, intra-renal, and post-renal causes (Hulse & Davies 2015). AKI has significant morbidity, mortality, and cost effects, and is sometimes preventable (Vrtis, 2013). Identifying individual RNs who significantly increase or decrease the probability of AKI would demonstrate the last point – that RNs can cause and prevent at least some AKI. Indeed, this finding is supported by our results.

## Methods

### Data and Panel Construction

We use data from the MIMIC-III database, version 1.4. MIMIC-III (Medical Information Mart for Intensive Care III) is a large, freely available database comprising de-identified health-related data associated with over forty thousand patients who stayed in critical care units of the Beth Israel Deaconess Medical Center between 2001 and 2012 (Johnson et al., 2016). The database contains data from two ICU medical records systems, Careview and Metavision, as well as data from the hospital laboratory system and the hospital EHR. Since the Metavision data is missing some RN notes, but the Careview data is not, we limit our analysis to ICU stays described by Careview data only.

We analyze single-service, uninterrupted, adult (>=18yo) ICU stays (n=26,656), summarized in Table 1. They are from the MICU, SICU, CCU, TSICU, and CSRU. Uninterrupted stays are not transferred to other ICU locations in the same stay, which MIMIC allows, and single-service stays do not change service lines, for example, from a surgery service to a medicine service, in the same ICU stay. Selecting single-service, uninterrupted stays helps preserve the fixed effect assumption, that each substay fixed effect is constant with constant effects in the substay. Abrupt changes in patient states accompanying stay-interruption would often violate this assumption for the substay in which the interruption occurs.

**Table 1.** Characteristics of adult ICU stays modeled

|  | Stays | LOS(d)* | Age* | AKI | LOS if AKI* | Age if AKI* |
| --- | --- | --- | --- | --- | --- | --- |
| All | 26,656 | (2.2, 4.1) | (65, 74) | 4,699 | (4.0, 7.0) | (67, 69) |
| MICU | 9,877 | (2.2, 4.2) | (64, 78) | 1,567 | (4.1, 6.8) | (63, 70) |
| SICU | 4,150 | (2.5, 5.0) | (63, 71) | 701 | (4.4, 8.5) | (60, 60) |
| CCU | 4,228 | (2.0, 3.4) | (70, 82) | 690 | (3.4, 4.9) | (74, 75) |
| TSICU | 3,035 | (2.0, 4.0) | (57, 65) | 399 | (5.7, 9.8) | (65, 73) |
| CSRU | 5,366 | (2.1, 3.7) | (67, 68) | 1,342 | (4.0, 6.9) | (69, 67) |

\*(median, mean)

### AKI detection, recovery, and timing

Computing AKI occurs in four broad steps: 1) detect AKI by absolute or relative changes in creatinine, 2) assign AKI to likely occurrence times, 3) remove redundant and borderline cases, and 4) compute AKI recovery in order to recognize subsequent AKI in the same stay.

We use the KDIGO criteria to detect AKI (Khwaja, 2012; Luo, 2014). Specifically, we apply the absolute creatinine (Cr) change criteria (increase in serum Cr by at least 0.3 mg/dl within 48h) to all hospitalization creatinine ordered pairs (Cr1, Cr2)_1_, where Cr2 succeeds Cr1 by less than 48h. Each pair that satisfies the absolute Cr change criteria (Cr2 > Cr1 + 0.3) is labeled a *precursor AKI type 1* (*prAKI*_*1*_) *detected* at time t_d_ when the specimen for Cr2 is collected.

Next we apply the relative change criteria of KDIGO to all creatinine ordered pairs (Cr1, Cr2)_2_ where Cr2 succeeds Cr1 within 7d. Each pair where Cr1 >= 1.5x Cr2 is labeled a *precursor AKI type 2* (*prAKI*_*2*_) *detected* at the time when the specimen for Cr1 was obtained. The relative change criteria use a future lower Cr as the baseline to detect AKI retrospectively, for example, on significant improvements in Cr after admission where the initial rise in Cr sometimes happens before Cr levels are obtained. MIMIC contains lab data for the entire hospitalization, and Cr1 and Cr2 can occur outside of the ICU stay. We account for this in detailed implementation logic.

Each AKI is detected after it occurs, and the rise in serum Cr concentrations after kidney injury follows complex kinetics. Models of these kinetics, however, show that after kidney injury, creatinine rises at a relatively constant rate of 0.5 mg/dl per 12 hours, regardless of baseline Cr (Waikar et al., 2009). For PrAKI_1_, this allows us to locate AKI *occurrence* based on differences in (Cr1, Cr2) and the time that separates them. PrAKI_1_ occurs at time t_o_ = t_d_ – (Cr2 – Cr1) / 0.5 * 12 h. Using this logic, most ICU PrAKI_1_ occur 6 to 18 hours before detection. For PrAKI_2,_ we take a different approach. Most PrAKI_2_ occur before a creatinine measurement is obtained. Therefore, we locate PrAKI_2_ to a default 12 hours before the specimen for Cr1 is collected. This has little effect, if any, on subsequent AKI calculations, all of which are based on AKI occurrence times, not detected times.

In the next step, we distill AKI from prAKI_1_ and prAKI_2_. First, we remove prAKI_1_ that have a subsequent Cr in the next 24h less than Cr1 + 0.3. This eliminates events caused by transiently elevated Cr that quickly return to non-elevated levels relative to Cr1. Next, for each prAKI_1_ and prAKI_2_ we compute *recovery*, if one exists, which occurs when creatinine first returns to levels at or below baseline (Formi et al., 2017). For prAKI_1,_ this happens when creatinine first returns to a level less than Cr1 + 0.3, and for prAKI_2_, this occurs at Cr2, since Cr2 is, by convention, the baseline for prAKI_2_. Any prAKI of either type that falls within the recovery period of another prAKI of either type is considered a continuation of a prior event and is removed. Remaining prAKI of either type are retained as AKI. AKI detection is binary. We do not attempt to stage AKI in this study.

In our implementation, additional AKI cannot occur during the recovery period of a preceding AKI, making recovery periods *AKI-ineligible*. Related to this concept is elevated Cr due to ongoing renal insufficiency due to chronic kidney injury (CKI). In our analysis, once patients have Cr greater than 4 mg/dl, they are AKI-ineligible until Cr returns to levels below 4. This prevents the detection of AKI on top of significant CKI, but we do this for two reasons - 1) Models of creatinine kinetics assume baseline creatinine is less than 4 mg/dl, and 2) in our experience, once Cr is above 4 mg/dl, Cr levels can be quite variable, often triggering AKI criteria even though there is no clear indication that AKI has occurred. AKI that cause Cr to first increase above 4 are detected by the criteria employed.

### Panel Construction and AKI-eligible segments

The MICIC III data are rich and contain many timed observations of patients throughout ICU stays (Johnson et al., 2016). To construct our panel, we begin by dividing each stay into contiguous, non-overlapping 3-hour segments and indexing them. For each segment, we compute a set of explanatory variables, an AKI indicator, and AKI eligibility. Then we work back in time from ICU discharge or AKI to construct 12-hour substays. Each 12-hour substay ends with the 3-hour segment that contains either AKI or discharge from the ICU, and the first substay at the beginning of each ICU stay is typically fewer than four segments. Substays cover contiguous AKI-eligible segments.

### Explanatory variables

Meaningful explanatory variables must be non-constant or have non-constant effects at the 12-hour substay, the length of each fixed effect interval. Amongst these, nursing assignment is of primary interest.

We define a *high-volume* RN as one with at least 100 nursing notes. Those with fewer than 100 notes are classified as *low-volume* RNs. RNs are assigned to 3-hour patient segments by matching time-labeled input events, output events, and vital signs to unique RN identifiers, then checking that the assignment has a matching note from the same RN. Specifically, an RN is assigned to a segment if an event in the segment has the RN identifier and the same RN writes a nursing note in the next 12 hours. This note-corroboration logic limits assignments to RNs with significant segment-level patient interactions. All individual RN assignments are binary at the segment level, and multiple RNs can be assigned to the same patient-segment.

For each 3-hour segment, we also compute a full panel of segment start-hour indicators, and interact them with ICU locations to capture any systematic time-of-day dependent features in each ICU. Finally, we compute cumulative low-volume RN assignment for each segment.

### Model

We construct a linear probability model with 12-hour substay-level fixed effects, and estimate it on AKI-eligible segments. The model is specified as follows.

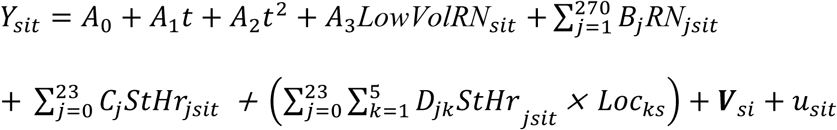

Y_s*it*_ is the probability of AKI in AKI-eligible segment *t* of patient stay *s*, where segment *t* resides in substay *i. StHr*_*jsit*_ is an indicator variable that records the clock hour in which segment *t* begins. Loc_js_ an indicator for the ICU location of stay *s*. Loc_ks_ by itself is a fixed effect, but its interaction with start hour is not, and accounts for any systematic differences in any time-of-day patterns between ICUs, like RN changeover times, MD rounding times, lab collection times, etc. *LowVolRN*_*s*it_ is the cumulative number of segments assigned to low-volume RNs in substay *i* through segment t. *RN*_j_ identifies a unique high-volume RN, and RN_jsit_ is the *cumulative* number of segments assigned to RN *j* in substay *i* through segment *t*. B_j_ is the marginal effect of RN *j* on Y_sit_. **V**_si_ is a vector of observed and unobserved variables that are constant in substay *i* of stay *s*, and u_sit_ is a random (idiosyncratic) error. We present results treating **V**_si_ as a vector of fixed effects and estimate a linear probability model with bootstrapped standard errors. We performed a Hausman test to confirm that fixed-effects estimation is preferred to random effects estimation (p<0.000).

We use a linear probability model (LPM) instead of a non-linear model for several reasons. First, despite their well-known limitations, LPMs almost always provide marginal estimated effects that are nearly as good as non-linear models, with similar standard errors (Wooldridge, 2010; Angrist & Pischke, 2013). Second, LPMs are much easier to estimate, especially on large panel data, and are easier to interpret since marginal effects are on the probability scale. Finally, the logistic fixed-effects estimator drops substays where the dependent variable does not change and would throw out AKI-eligible segments in substays where AKI does not occur. The LPM does not have these problems, and several well-known experts have advocated for their use when the primary goal of analysis is to estimate the marginal effects of explanatory variables (Angrist & Pischke, 2013; Wooldridge, 2010).

All analysis was done using STATA v16 (StataCorp. 2019. *Stata Statistical Software: Release 16*. College Station, TX: StataCorp LLC.) Investigators were certified to use the MIMIC-III database. All data were de-identified and no additional IRB approval was required.

## Results

ICU stays (n=26,656) have mean and median total ICU length of stay (LOS) of 4.1 days and 2.2 days, respectively. In 4,154 of these stays, 4,699 AKI were detected. Stays with at least one AKI have mean and median total LOS of 7.4 and 3.8 days, respectively (Table 1). Stays comprise 900,181 3-hour segments, 71% of which are AKI-eligible. There are 170,983 substays, each comprised of one or more contiguous AKI-eligible segments. Each has a fixed effect in the model. From these, the RN_j_ effects, B_j_, are of primary interest (Table 2). Each B_j_ estimates the attributable marginal effect of an RN_j_ segment-level assignment on the segment-level probability of AKI.

**Table 2.**
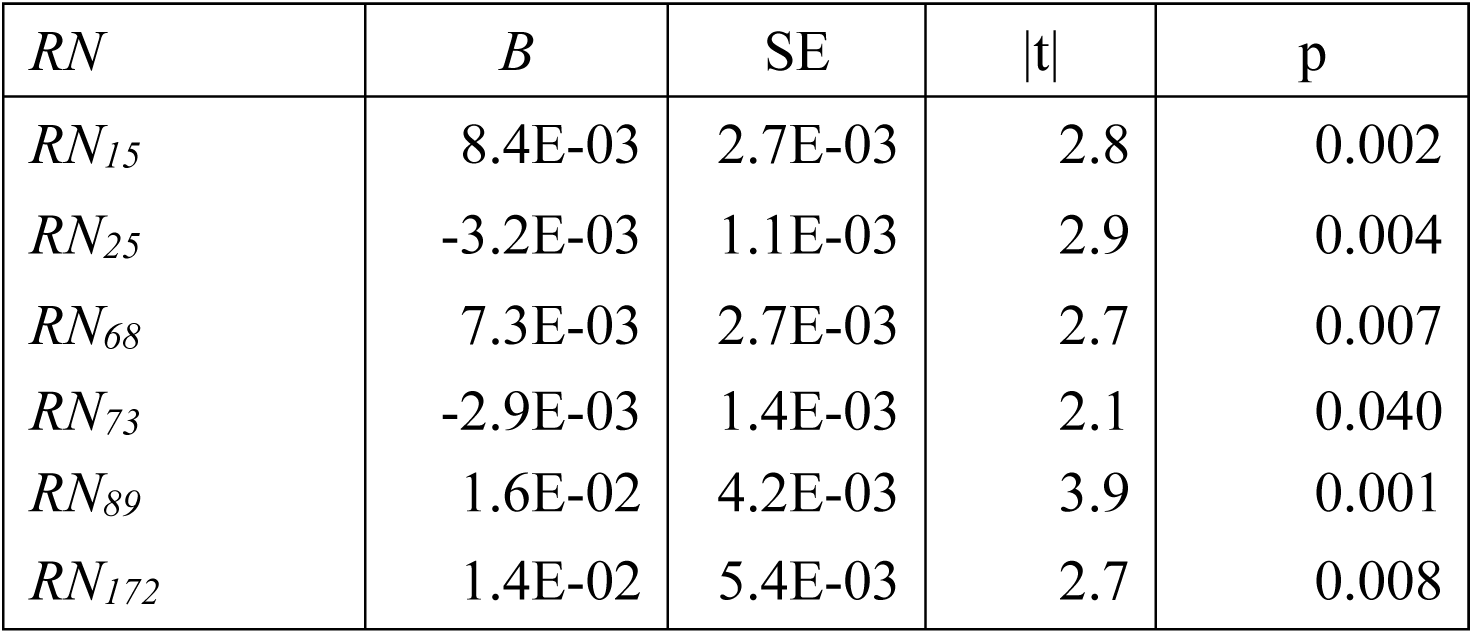
Select outlier RN

We test each B_j_ against the null hypothesis H_0_: B_j_ = 0, i.e. no effect, and identify 2 standard error (SE) outliers. Since there are 270 RNs, we expect to find about 14 outliers by chance, 7 high and 7 low. We find 33: 22 high-side outliers, and 11 low-side outliers. Outlier B_j_ range from about 0.4 to 2.2 (mean = 0.96) percentage points on the high side, and −0.6 to −0.3 (mean = −0.43) percentage points on the low side. Each outlier RN works primarily in one ICU. Individual RN effects, by ICU primarily worked, are shown in Figure 1.

**Figure 1:**
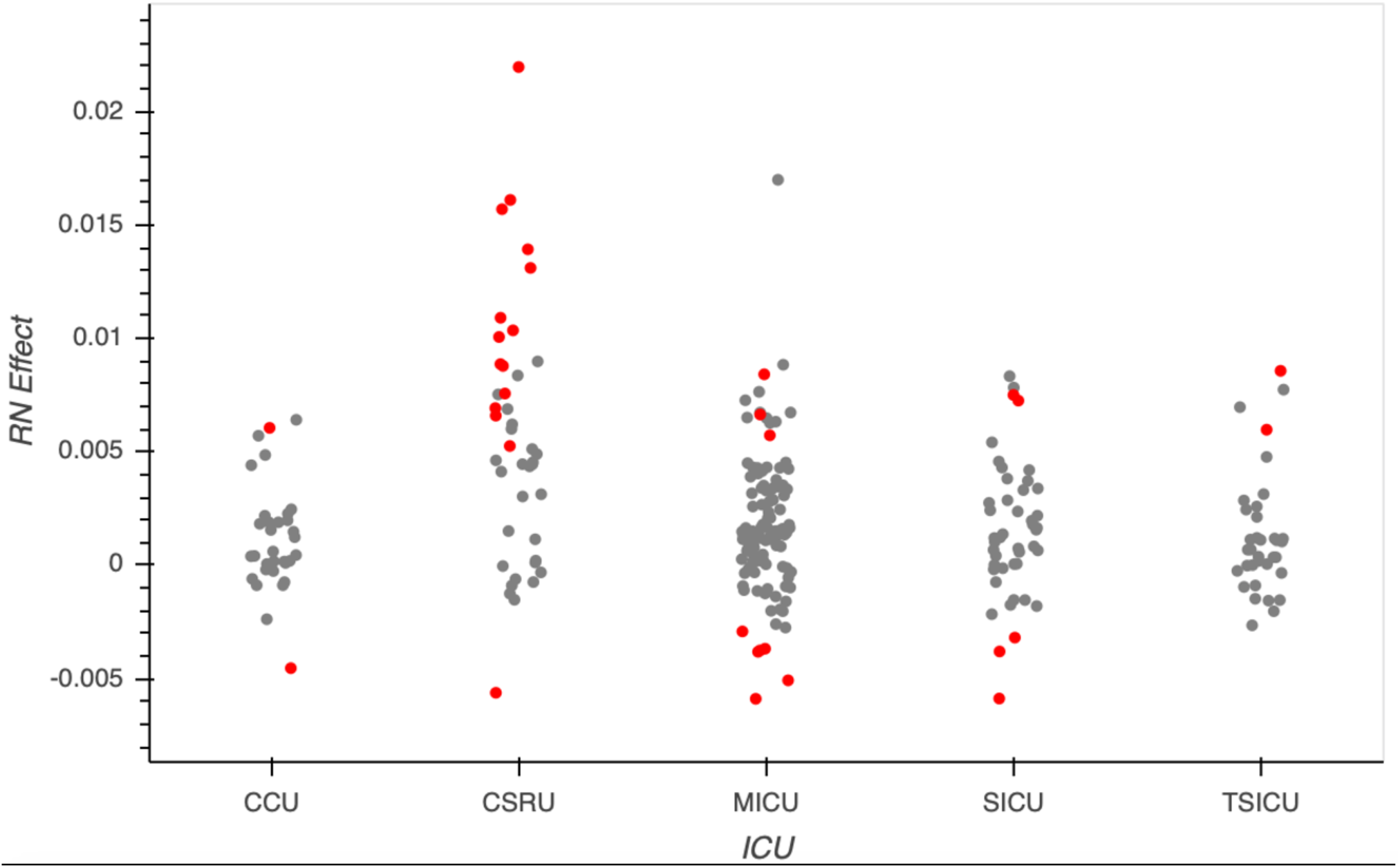
Individual RN effects (model outliers in red)

We obtain the substay-level effect and the work-shift effect of RN *j* as follows. First, we note that a 12-hour work shift by RN *j* spans 4 contiguous one-patient segments. If RN *j* is assigned to an average of 1.5 patients at a time, on average, then in one 12-hour work shift, RN *j* is assigned to 6 patient-segments. Of these, about 71% are AKI-eligible, on average, so the average work-shift effect of RN j is about 4.2B_j_. Using this estimate, each high-side outlier RN increases the probability of AKI by about 4 percentage points per work shift and is associated with about one extra AKI per 25 work-shifts. Each low-side outlier reduces the probability AKI by about 2 percentage points per work shift and is associated with about 1 fewer AKI per 50 work shifts.

Other significant explanatory variables (p<0.01) include *t*, all locations and their *t* interactions, *StHr* 10-13 and 18-20, and a number of *StHr* location interactions.

## Discussion

While it is widely believed that nursing affects outcomes (McHugh et al, 2013; Lake et al, 2012; Dall et al, 2009; Blegen, 2013; Dabney 2015; Konetzka 2007; Kutney-Lee 2015; Mchugh 2013; Needleman 2002; Weiss 2011; Yakusheva 2014), estimating the effects of individual nurses on outcomes has been elusive. In this work, we present a model that estimates the effects of individual RNs on the probability of a significant clinical event, AKI in the ICU. Using substay-level fixed effects, the model accounts for significant sources of unobserved heterogeneity and produces estimates of nursing effects that we believe are both conservative and approximately causal. This implies that some AKI are nursing-linked and are preventable. With caution, our model could be used to estimate additional AKI caused, as well as those prevented, and attribute them to individual nurses. Each high-side RN outlier causes about one additional AKI per 25 12-hour work shifts, and each low-side RN outlier prevents about 1 AKI in 50 work shifts. The number of outliers, high and low, depends on the critical value cutoff used. The model generated 22 high-side outliers and 11 low-side outliers at 2SE cutoffs, for each type, well in excess of the numbers of expected by chance. Since the model assumes that marginal RN assignment effects are confined to the substay, we believe these are conservative estimates of the number of additional AKI caused and prevented.

We chose AKI because it is a readily computable, common, clinically significant, and costly outcome associated with a number of other important hospital complications including volume depletion, sepsis, drug toxicity, and myocardial infarction (Alscher et al, 2019). Furthermore, nurses have a key role in preventing and treating AKI (Vrtis, 2015; Aitken et. al. 2013; Hulse & Davies, 2015).

Our work is most closely related to that of Yakusheva, et. al. who estimates the value-added effects of individual nurses over a subset of adult inpatient stays (Yakusheva, Lindrooth, & Weiss, 2014). Yakusheva’s work is impressive and important and is the first to our knowledge that attempts to estimate the marginal effects of individual nurses on patient outcomes. It correctly recognizes the central problem of non-random nursing assignment and attempts to control for it by adding a number of explanatory variables including patient demographics, insurance type, unit assignment, diagnosis, medical team assignments, and calendar week and day indicators. The approach appears to control for initial non-random assignment, but may be limited in its ability to control for ongoing non-random assignment after admission. The model includes a variable for each high-volume nurse, like ours, and estimates the *binary* assignment effects of individual nurses on several admission-level outcomes. Marginal individual nurse effects are adjusted by subtracting off the average nurse effect, and the resulting value-add effects are arguably conservative.

Our work differs in from Yakusheva in several ways. First, instead of estimating the binary effect of individual nurse-to-patient assignment on admit-level outcomes, we model the short-term effects of individual RN exposures (in 3-hour segments) on a specific clinical outcome, AKI in the ICU. Second, we construct a segment-in-substay panel and use the fixed-effects estimator to control for unobserved heterogeneity at the substay level, which incrementally adjusts for complex observed and unobserved factors associated with non-random nursing assignment, both on admission, and throughout the ICU stay. Third, while Yakusheva uses the term “fixed effect” to refer to observed, model-included variables that are constant in patient-unit-nurse observations, we use the term in the spirit of unobserved effects models, where repeated segment-level observations in substays provide a panel for the fixed-effects estimator to demean changing variables like cumulative individual RN assignments, and control for all substay-constant variables correlated with them, whether observed or not. While this approach does not allow us to compute estimates for the fixed effects directly, it robustly controls for unobserved heterogeneity that cannot always be captured by observable explanatory variables. Therefore, the fixed-effect estimator likely produces relatively unbiased, causal estimates of the effects of individual nurses, our primary interest.

Our work is tangentially related to evaluations of teacher value and the specification of education production functions found in the education economics literature. Some of these studies employ fixed effects to control for unobserved heterogeneity in individual students (Dynarsky et. al., 2017), which is similar to our use of fixed effects to control for unobserved heterogeneity in patient-specific ICU substays.

We envision several applications for this work. They include targeted outcome-related education to nurses, performance monitoring to detect nursing burnout, and incentives targeting to reward and retain the high-performing nurses.

Limitations of our work include those of the fixed-effects estimator, primarily the inability to compute estimates of the fixed effects themselves. These include the effects on the probability of AKI produced by specific clinical conditions, unit assignments, and nursing characteristics like education, experience, and tenure. However, those are not of primary interest in this study. Other potential limitations are associated with the granularity of the 12-hour substay. While many features are constant over the substay, others like clinical state, are only approximately so. Within-substay changes to clinical state may affect nursing assignments in the substay, especially in ICU stays with care discontinuities like changing ICU locations and changing medical/surgical services, which we excluded. We do not believe that additional unobserved variation in-substay is a significant concern, and that most non-random assignment preferences are well handled by the substay fixed effects.

Despite these limitations, we believe that our approach provides relatively unbiased estimates of individual nursing effects on the likelihood of AKI in the ICU. To our knowledge, this is the first example in the literature to estimate the effects of individual nurses on a significant clinical outcome.

## Conclusion

Nursing is the largest labor force in healthcare. However, quantifying the effects of individual nurses on clinical outcomes has been elusive. We have introduced a new framework that estimates the effects of individual RNs on acute kidney injury in the ICU. To our knowledge, this is the first work to employ fine-grained fixed effects to control for unobserved heterogeneity correlated with nursing assignment. We show that some AKI are nursing-linked, and that estimates of additional AKI caused, as well as those prevented, can be assigned to individual nurses. We believe this a significant contribution to the healthcare economics and policy literature and hope that it encourages additional investigations.

## Data Availability

All data analyzed in this manuscript is available here: https://mimic.physionet.org

## Conflict of interest

All authors have a financial interest in Indicator Sciences, LLC

## Funding sources

Indicator Sciences, LLC

## IRB

All data used were de-identified and publicly available. No IRB approval was required.

## Intellectual property notice

Some methods and related aspects described in this paper are the intellectual property of Indicator Sciences, LLC and are patent pending.

## Notes

### Competing Interest Statement

All authors are employees of Indicator Sciences LLC which funded this work.

### Funding Statement

All authors are employees of Indicator Sciences LLC which funded this work.

### Author Declarations

All analysis was performed on the MIMIC-III database, a publicly available, de-identified clinical database. IRB approval was not required.

## References

Aiken, L. H. (2003). Educational Levels of Hospital Nurses and Surgical Patient Mortality. JAMA: The Journal of the American Medical Association, 290(12), 1617–1623. doi:10.1001/jama.290.12.1617

Aiken, L. H., Cimiotti, J. P., Sloane, D. M., Smith, H. L., Flynn, L., & Neff, D. F. (2011). Effects of Nurse Staffing and Nurse Education on Patient Deaths in Hospitals With Different Nurse Work Environments. Medical Care, 49(12), 1047–1053. doi:10.1097/mlr.0b013e3182330b6e

Aitken, E., Carruthers, C., Gall, L., Kerr, L., Geddes, C., & Kingsmore, D. (2013). Acute kidney injury: Outcomes and quality of care. Qjm, 106(4), 323–332. doi:10.1093/qjmed/hcs237

AKI, Epidemiology of. (n.d.). SpringerReference. doi:10.1007/springerreference_224911

Alscher, M. D., Erley, C., & Kuhlmann, M. K. (2019). Acute renal failure of nosocomial origin. Deutsches Aerzteblatt Online. doi:10.3238/arztebl.2019.0149

Angrist, J. D., & Pischke, J. (2013). 3.4.2/Limited Dependent Variables and Marginal Effects. In Mostly harmless econometrics: An empiricists companion. United States, NJ: Content Technologies.

Blegen, M. A., Goode, C. J., Park, S. H., Vaughn, T., & Spetz, J. (2013). Baccalaureate Education in Nursing and Patient Outcomes. JONA: The Journal of Nursing Administration, 43(2), 89–94. doi:10.1097/nna.0b013e31827f2028

Clarke, S. P., & Aiken, L. H. (2003). Failure To Rescue. AJN, American Journal of Nursing, 103(9), 13. doi:10.1097/00000446-200309000-00004

Dabney, B. W., & Kalisch, B. J. (2015). Nurse Staffing Levels and Patient-Reported Missed Nursing Care. Journal of Nursing Care Quality, 30(4), 306–312. doi:10.1097/ncq.0000000000000123

Dall, T. M., Chen, Y. J., Seifert, R. F., Maddox, P. J., & Hogan, P. F. (2009). The Economic Value of Professional Nursing. Medical Care, 47(1), 97–104. doi:10.1097/mlr.0b013e3181844da8

Formi, L. G, Darmon, M, Ostermann, M. Oudemans-van Straaten, H. M, Pettilä, V., Prowle, J. R. Schetz, M. & Joannidis, M. (2017) Renal Recovery After Acute Kidney Injury. Intensive Care Med, 43(1), 855–866. doi:10.1007/s00134-017-4809-x

Hoste, E. A. (2009). Epidemiology of Nosocomial Acute Kidney Injury. Critical Care Nephrology, 86–91. doi:10.1016/b978-1-4160-4252-5.50018-6

Johnson, A. E., Pollard, T. J., Shen, L., Lehman, L. H., Feng, M., Ghassemi, M., … Mark, R. G. (2016). MIMIC-III, a freely accessible critical care database. Scientific Data, 3(1). doi:10.1038/sdata.2016.35

Konetzka, R. T., Stearns, S. C., & Park, J. (2007). The Staffing-Outcomes Relationship in Nursing Homes. Health Services Research, 43(3), 1025–1042. doi:10.1111/j.1475-6773.2007.00803.x

Kutney-Lee, A., Stimpfel, A. W., Sloane, D. M., Cimiotti, J. P., Quinn, L. W., & Aiken, L. H. (2015). Changes in Patient and Nurse Outcomes Associated With Magnet Hospital Recognition. Medical Care, 53(6), 550–557. doi:10.1097/mlr.0000000000000355

Khwaja, A. (2012). KDIGO Clinical Practice Guidelines for Acute Kidney Injury. Nephron, 120(4), C179–C184. doi:10.1159/000339789

Lake, E.T., Staiger D., Horbar J., et al. (2012) Association Between Hospital Recognition for Nursing Excellence and Outcomes of Very Low-Birth-Weight Infants. JAMA, 307(16),1709–1716. doi:10.1001/jama.2012.504

Lameire, N. (2018). Prevention of acute kidney injury. Oxford Medicine Online. doi:10.1093/med/9780199592548.003.0224_update_001

Luo, X., Jiang, L., Du, B., Wen, Y., Wang, M., & Xi, X. (2014). A comparison of different diagnostic criteria of acute kidney injury in critically ill patients. Critical Care, 18(4). doi:10.1186/cc13977

Mchugh, M. D., Kelly, L. A., Smith, H. L., Wu, E. S., Vanak, J. M., & Aiken, L. H. (2013). Lower Mortality in Magnet Hospitals. Medical Care, 51(5), 382–388. doi:10.1097/mlr.0b013e3182726cc5

Mehta, R. L., Kellum, J. A., Shah, S. V., Molitoris, B. A., Ronco, C., Warnock, D. G., & Levin, A. (2007). Acute Kidney Injury Network: Report of an initiative to improve outcomes in acute kidney injury. Critical Care, 11(2). doi:10.1186/cc5713

Murnane, R. J. 1975. The Impact of School Resources on the Learning of Inner City Children. Cambridge, MA: Ballinger Publishing Co.

Needleman, J., Buerhaus, P., Mattke, S., Stewart, M., & Zelevinsky, K. (2002). Nurse-Staffing Levels and the Quality of Care in Hospitals. New England Journal of Medicine, 346(22), 1715–1722. doi:10.1056/nejmsa012247

Ostermann, M. (2018). Epidemiology, Incidence, Risk Factors, and Outcomes of Acute Kidney Injury. Core Concepts in Acute Kidney Injury, 3–11. doi:10.1007/978-1-4939-8628-6_1

Park, J., & Stearns, S. C. (2009). Effects of State Minimum Staffing Standards on Nursing Home Staffing and Quality of Care. Health Services Research, 44(1), 56–78. doi:10.1111/j.1475-6773.2008.00906.x

Scott, D. W. (1979). On optimal and data-based histograms. Biometrika, 66(3), 605–610. doi:10.1093/biomet/66.3.605

Vrtis, M. C. (2013). Preventing and Responding to Acute Kidney Injury. AJN, American Journal of Nursing, 113(4), 38. doi:10.1097/01.naj.0000428738.33180.2f

Waikar, S. S., & Bonventre, J. V. (2009). Creatinine Kinetics and the Definition of Acute Kidney Injury. Journal of the American Society of Nephrology, 20(3), 672–679. doi:10.1681/asn.2008070669

Welton, J. M. (2015). Hospital Nursing Workforce Costs, Wages, Occupational Mix,and Resource Utilization. JONA: The Journal of Nursing Administration, 45(Supplement), S10–S15. doi:10.1097/nna.0000000000000247

Weiss, M. E., O. Yakusheva, and K. L. Bobay. 2011. “Nursing Staffing, Readiness for Hospital Discharge, and Post-Discharge Utilization.” Health Services Research 46 (5): 1473–94.

Wooldridge, J. M. (2010). 15.2/Linear Probability Model for Binary Response. In Econometric analysis of cross section and panel data (2nd ed.). Cambridge, MA, MA: MIT Press.

Yakusheva, O., Lindrooth, R., & Weiss, M. (2014). Nurse Value-Added and Patient Outcomes in Acute Care. Health Services Research. doi:10.1111/1475-6773.12236

